# Deep learning model for prenatal congenital heart disease (CHD) screening generalizes to the community setting and outperforms clinical detection

**DOI:** 10.1101/2023.03.10.23287134

**Authors:** Chinmayee Athalye, Amber van Nisselrooij, Sara Rizvi, Monique Haak, Anita J. Moon-Grady, Rima Arnaout

## Abstract

**Objective:** Congenital heart defects (CHD) are still missed despite nearly universal prenatal ultrasound screening programs, which may result in severe morbidity or even death. Deep machine learning (DL) can automate image recognition from ultrasound. The aim of this study was to apply a previously developed DL model trained on images from a tertiary center, to fetal ultrasound images obtained during the second-trimester standard anomaly scan in a low-risk population.

**Methods:** All pregnancies with isolated severe CHD in the Northwestern region of the Netherlands between 2015 and 2016 with available stored images were evaluated, as well as a sample of normal fetuses’ examinations from the same region. We compared initial clinical diagnostic accuracy (made in real time), model accuracy, and performance of blinded human experts with access only to the stored images (like the model). We analyzed performance by study characteristics such as duration, quality (independently scored by study investigators), number of stored images, and availability of screening views.

**Results:** A total of 42 normal fetuses and 66 cases of isolated CHD at birth were analyzed. Of the abnormal cases, 31 were missed and 35 were detected at the time of the clinical anatomy scan (sensitivity 53 percent). Model sensitivity and specificity was 91 and 93 percent, respectively. Blinded human experts (n=3) achieved sensitivity and specificity of 55±10 percent (range 47-67 percent) and 71±13 percent (range 57-83 percent), respectively. There was a statistically significant difference in model correctness by expert-grader quality score (p=0.04). Abnormal cases included 19 lesions the model had not encountered in its training; the model’s performance (15/19 correct) was not statistically significantly different on previously encountered vs. never before seen lesions (p=0.07).

**Conclusions:** A previously trained DL algorithm out-performed human experts in detecting CHD in a cohort in which over 50 percent of CHD cases were initially missed clinically. Notably, the DL algorithm performed well on community-acquired images in a low-risk population, including lesions it had not been previously exposed to. Furthermore, when both the model and blinded human experts had access to stored images alone, the model outperformed expert humans. Together, these findings support the proposition that use of DL models can improve prenatal detection of CHD.

## Introduction

Congenital heart disease (CHD) is the most common birth defect but is nevertheless rare, affecting ∼1 percent of births per year(1,2). Prenatal diagnosis of CHD reduces morbidity and mortality in neonates and increases therapeutic options, including *in utero* interventions.

Second-trimester ultrasound is universally recommended due to its potential to recognize 90 percent of severe congenital heart disease(3,4). However, in practice as little as 30-50 percent are detected(5). This gap in detection is hypothesized to be due to poor image quality and inability to recognize abnormal images and is higher outside of expert centers(5). Quality improvement programs can increase detection(6–9), but cannot be applied and sustained universally(5). Therefore, automated, scalable, and robust approaches to prenatal CHD screening are needed.

Previously(10), we showed that a deep learning (DL) model could be used to detect CHD. DL, a form of artificial intelligence(11), is a computational method which has the potential for automated and scalable image analysis(12). Testing this model in two tertiary care centers performed well.

However, for DL to truly democratize accurate prenatal CHD detection(13), it must also perform well in the community. Community imaging may differ from that of tertiary centers: for example, scanning expertise may be lower, captured images may be fewer and may be stored in low-resolution formats. Scanning protocols may capture different screening views(4,14) and may even vary by sonographer(15). Finally, the patient population may vary with respect to CHD prevalence, body habitus, and other factors.

Furthermore, an optimal DL-based fetal screening tool should provide explainability. Our DL model was designed such that model-learned image features correlated to relevant screening views and important anatomic structures within those views(10), inviting analysis of model performance along these features. However, few community imaging cohorts with these types of annotations exist. In van Nisselrooij et al 2020, screening fetal ultrasound for all complex CHD births in 2015-2016 in the northwestern Netherlands, were collected and graded as to the completeness and quality of views obtained(16), providing an excellent opportunity to test DL model performance in a well-phenotyped community imaging cohort. We hypothesized the DL model could be applied successfully to community-based screening cohort.

## Methods

### Datasets

Pregnancies affected by severe congenital heart defects, defined as the need for surgery in the first year of life, were extracted from the PRECOR registry(14,17). All cases in PRECOR with births in 2015 and 2016, both detected and not detected, were asked for consent to collect their images from the second trimester anomaly scan at the initial community-based screening facility(16). To be able to test the algorithm on images from normal pregnancies as well, we additionally collected images of pregnancies without cardiac or other birth defects from the same screening facilities in the same region during the same time period. We randomly selected women from local imaging databases to consent (see Ethics and approvals).

### Expert grading

Non-blinded expert grading of study quality: image quality assessment for both CHD and normal cases was performed as previously described(16). Briefly, each cardiac view that was standard in the Netherlands at the time—3VV, right ventricular outflow tract (RVOT), LVOT, and 4CV— was scored for completeness and technical correctness on a scale of 0 to 5 by two fetal echocardiography experts. The experts also measured study duration in minutes, total number of stored images per study, and graded fetal position, amniotic fluid, image quality, and magnification. Finally, they determined whether or not the CHD was discernible from the stored imaging.

Non-blinded view sorting: to obtain ground-truth labels for view sorting, an expert fetal cardiologist labeled each image frame by view, as previously described(10).

Blinded diagnosis of normal vs CHD by clinicians based only on stored images: human fetal cardiac experts blinded to the diagnosis, clinical impression, composition (percent abnormal) of the dataset, and purpose of the study served as the human study subjects. They viewed the stored images and graded each heart as normal vs CHD. Human subjects had access to both color doppler and grayscale images and videos (Table S1) in their native format (.jpg for still images and .avi for video). Years of fetal cardiac ultrasound experience ranged from 15 to greater than 25 years.

### Model inference and screening diagnosis

Clinical images in .jpg (still image) and .avi (video) formats were de-identified and converted to .png as previously described(10). As before, only grayscale images were used (Table S1). Images were input into the DL model as previously described(10) with the following modifications to the aggregation of predictions on individual images into an overall study-level decision: First, due to the difference between the Netherlands national scanning protocol at that time and International Society of Ultrasound in Obstetrics and Gynecology (ISUOG) protocol(4) (Fig. 1), only the three vessel view (3VV), left ventricular outflow tract (LVOT), and four-chamber view (4CV) views were utilized for CHD detection(18). Second, a slightly different method was used to filter low-quality predictions. A neural network’s classification on a particular image is actually a set of probabilities of the image belonging to each of the possible classes (the softmax vector); by default, the image is assigned to the class with the highest probability. Previously, only images where the highest prediction probability was at or above the first quartile of probabilities for that view were deemed sufficient quality. Here, entropy across all six view predictions per image was calculated(19), where high entropy indicates model confusion between view categories – these images were discarded as low-quality and not used in the next step of predicting normal vs CHD. (iii) Similarly, for normal vs abnormal prediction for each image, an entropy threshold of 0.85 was used, corresponding to the model being at least 70 percent sure of its classification decision. Gradient-weighted class activation maps (GradCAMs) were computed on test images according to standard techniques(20).

**Figure 1.**
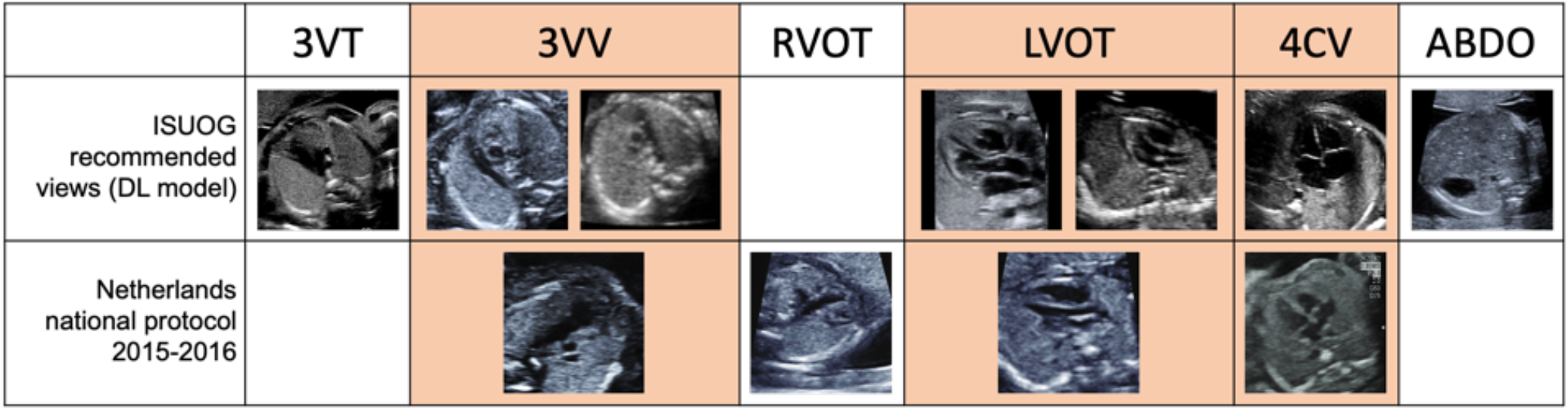
Differences in view acquisition protocol between development dataset and current cohort. The views in orange represent views that are common between the ISUOG and the Netherlands national protocols, and represent the views used for CHD detection in this study. Of note, our model was trained to recognize both 3VV and RVOT views as 3VV, and to recognize axial and sagittal LVOT views. 3VT, three-vessel-trachea; 3VV, three-vessel view; RVOT, right ventricular outflow tract; LVOT, left ventricular outflow tract; 4CV, four-chamber view; ABDO, abdomen. Netherlands national protocol images reprinted with permission from A.E.L. van Nisselrooij and M.C. Haak and with permission from Wiley.

### Statistical testing

Except where otherwise specified, the Mann-Whitney U test was used for all statistical tests.

### Ethics and approvals

All investigations were performed in accordance with relevant national guidelines and regulations. All experimental protocols were approved by local institutional review/ethics committees. Participation of clinical experts as human subjects was deemed exempt research by the UCSF IRB. The Ethics Board of LUMC approved collection and analysis of images (IRB number P15.374), with written informed consent obtained from all subjects.

## Results

### Study cohort characteristics

The test dataset included 108 ultrasound studies from patients from 18 to 22 weeks of gestational age, comprising normal hearts and a range of CHD lesions as previously described(16) and in Table 1. Average number of items (still images or cine, grayscale and color) per study were 41±18 (range 6-103) and was not statistically different between normal and CHD cases (one sample t-test p=0.44). The DL model operates on grayscale images only (Table S1); average number of grayscale items per study was 35±18 (range 2-78), therefore the DL model had fewer stored items on which to make its decision (one-sample t-test between all items and grayscale items, p<0.01). Ten CHD studies had cine videos stored, while no normal studies had cine.

**Table 1.** CHD lesions present in the Netherlands cohort.

| **Lesion** | **Number**<br>**(percent)**<br>**of cases** | **Number correct,**<br>**clinical**<br>**diagnosis** | **Lesion included**<br>**in model**<br>**training** | **Number**<br>**correct,**<br>**model** |
| --- | --- | --- | --- | --- |
| **Normal** | 42 (39) | 42 | yes | 32 |
| **ALCAPA** | 1 (1) | 0 | no | 0 |
| **Aortic stenosis** | 4 (4) | 1 | yes | 2 |
| **Aortopulmonary**<br>**window** | 1 (1) | 0 | no | 1 |
| **AVSD** | 7 (6) | 5 | yes | 5 |
| **Coarctation of aorta** | 11 (10) | 3 | yes | 6 |
| **DORV** | 4 (4) | 4 | yes | 2 |
| **Double arch** | 2 (2) | 0 | no | 1 |
| **D-TGA** | 12 (11) | 11 | yes | 6 |
| **Interrupted aortic arch** | 1 (1) | 1 | no | 1 |
| **L-TGA** | 1 (1) | 1 | yes | 0 |
| **PAIVS** | 1 (1) | 1 | yes | 1 |
| **Pulmonary sling** | 1 (1) | 0 | no | 1 |
| **Pulmonic stenosis** | 1 (1) | 0 | yes | 1 |
| **RAI** | 1 (1) | 1 | yes | 1 |
| **Shone complex** | 2 (2) | 2 | no | 2 |
| **TAPVR** | 2 (2) | 0 | yes | 1 |
| **Tetralogy of Fallot** | 2 (2) | 1 | yes | 2 |
| **Truncus arteriosus** | 1 (1) | 1 | yes | 1 |
| **VSD** | 11 (10) | 2 | no | 9 |

ALCAPA, anomalous left coronary artery from the pulmonary artery; AVSD, atrioventricular septal defect; DORV, double outlet right ventricle; D-TGA, dextro-transposition of the great arteries; L-TGA, levo-transposition of the great arteries; PAIVS, pulmonary atresia with intact ventricular septum; RAI, right atrial isomerism; TAPVR, total anomalous pulmonary venous return; VSD, ventricular septal defect.

### Overall performance

In the test dataset, the sensitivity and specificity of the clinical diagnosis (“clinical sensitivity and specificity”) were 0.53 and 1.0, respectively. (Of note, these are not the same as the overall clinical sensitivity and specificity in the northwestern Netherlands(14) due to dataset construction, see Methods.) In contrast, the DL model’s sensitivity and specificity on stored grayscale images from this dataset were 91 and 93 percent, respectively. The model was able to grade 106 of the 108 studies; two studies were not graded by the model due to insufficient image data. Finally, the blinded clinical experts with access to all stored clinical images had a sensitivity of 55±10 percent (range 47-67 percent) and a specificity of 71±13 percent (range 57-83 percent).

Clinical sensitivity was statistically similar to that of the blinded experts (p-value 0.76, one-sample t-test), while model sensitivity was higher than the blinded experts (p-value 0.03, one-sample t-test). Model specificity is statistically similar to that of the blinded experts (p-value 0.11, one-sample t-test). (Note that because clinically normal studies were explicitly chosen in construction of the test dataset, a comparison of specificity between clinical diagnosis and blinded experts is less relevant).

### Performance on CHD cases

66 studies had CHD, 35 of which were initially detected clinically and 31 of which were missed. Expert non-blinded retrospective grading determined that of the 31 misses, 10 were evident on imaging but not recognized, 14 were due to imaging of low technical quality, and seven were considered inevitable based on stored imaging and despite adequate imaging quality. Of the CHDs missed clinically, the model detected an anomaly in 18/31 cases, including five of the seven deemed to be inevitable, while blinded clinicians detected 11±5 missed cases (range 8-17 of 31) including 0-2 of the seven inevitable cases. Of 42 clinically confirmed normals, the model got 32 correct, while blinded clinicians detected 32±4. These data are summarized in Table 2.

**Table 2.** Performance of clinical detection, deep learning model, and blinded clinical experts.

| **Cases (No.)** | **Clinical diagnosis, No. Correct** | **Non-blinded expert assessment (No.)** | **Model decision, No. Correct** | **Blinded clinicians, No. Correct avg±stdv** |
| --- | --- | --- | --- | --- |
| **Normal (42)** | 42 | — | 32 | 32±4 |
| **Abnormal, clinically correct (35)** | 35 | — | 24 | 25±3 |
| **Abnormal, clinically missed (31)** | 0 | Not recognized (10) | 4 | 4±2 |
| | | Technically poor imaging (14) | 9 | 6±4 |
| | | Inevitable (7) | 5 | 1±1 |
| **Overall sensitivity** | 53% | — | 91% | 55±10% |
| **Overall specificity** | 100% | — | 93% | 71±13% |

Of the 66 CHD cases, 47 were lesion types the model had encountered during its training (Table 1), while 19 cases represented lesions the model had not encountered during training, namely, anomalous left coronary artery from the pulmonary artery (ALCAPA), aortopulmonary window, double arch, interrupted aortic arch, pulmonary sling, Shone complex, and ventricular septal defect (VSD). The model detected 15 of these 19 cases.

Where the model classifies a particular image, we can visualize the areas in the image most important to the model’s decision using an algorithm called gradient-weighted class activation maps (GradCAM). For several CHD lesions, per-image prediction of normal vs not was largely consistent with clinical knowledge about which views are abnormal in a given lesion (Figs. 2-3). GradCAMs often, but not always, corresponded to anatomical structures of interest (Figs. 2-4). Additionally, we examined failure modes in model prediction, which can include errors at both view classification and normal/abnormal detection steps (Fig. S1); some of these per-image errors do not result in an overall incorrect prediction at a subject level.

**Figure 2.**
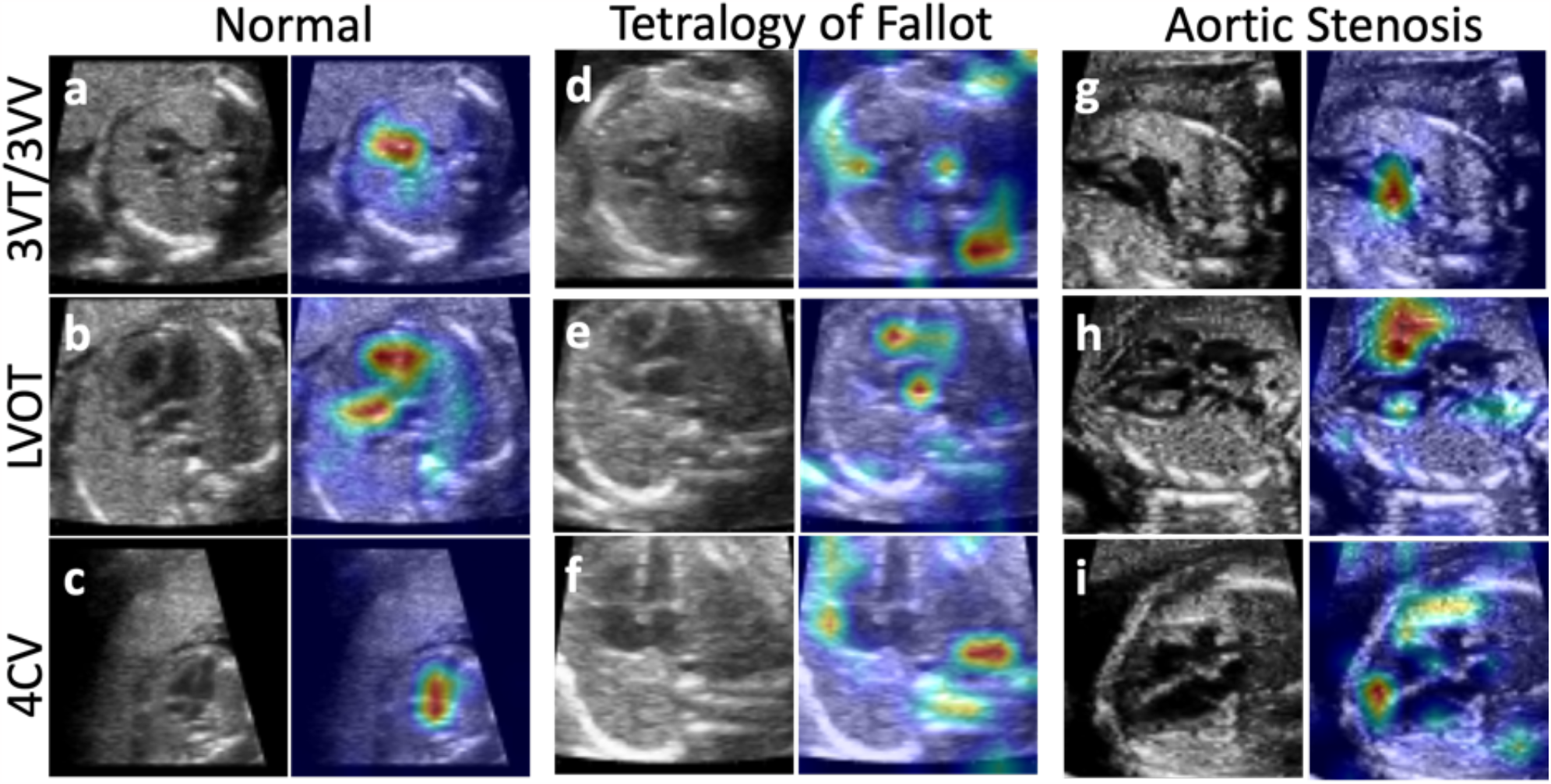
Diagnostic classifier in studies the model got correct and blinded experts missed. Model-labeled views (grayscale) with corresponding GradCAM images representing a heat map showing the areas of the image most important in model decision making. The model correctly identified the normal views (a-c), the abnormal 3VT and LVOT (d, e) and abnormal 4CV cardiac axis (f) in tetralogy of Fallot, and abnormal LVOT in aortic stenosis (h). The human experts misclassified these tetralogy and aortic stenosis patients as “normal”. Abbreviations: 3VT, three-vessels-and-trachea view; 3VV, three-vessel view; 4CV, four-chamber view; LVOT, left ventricular outflow tract.

**Figure 3.**
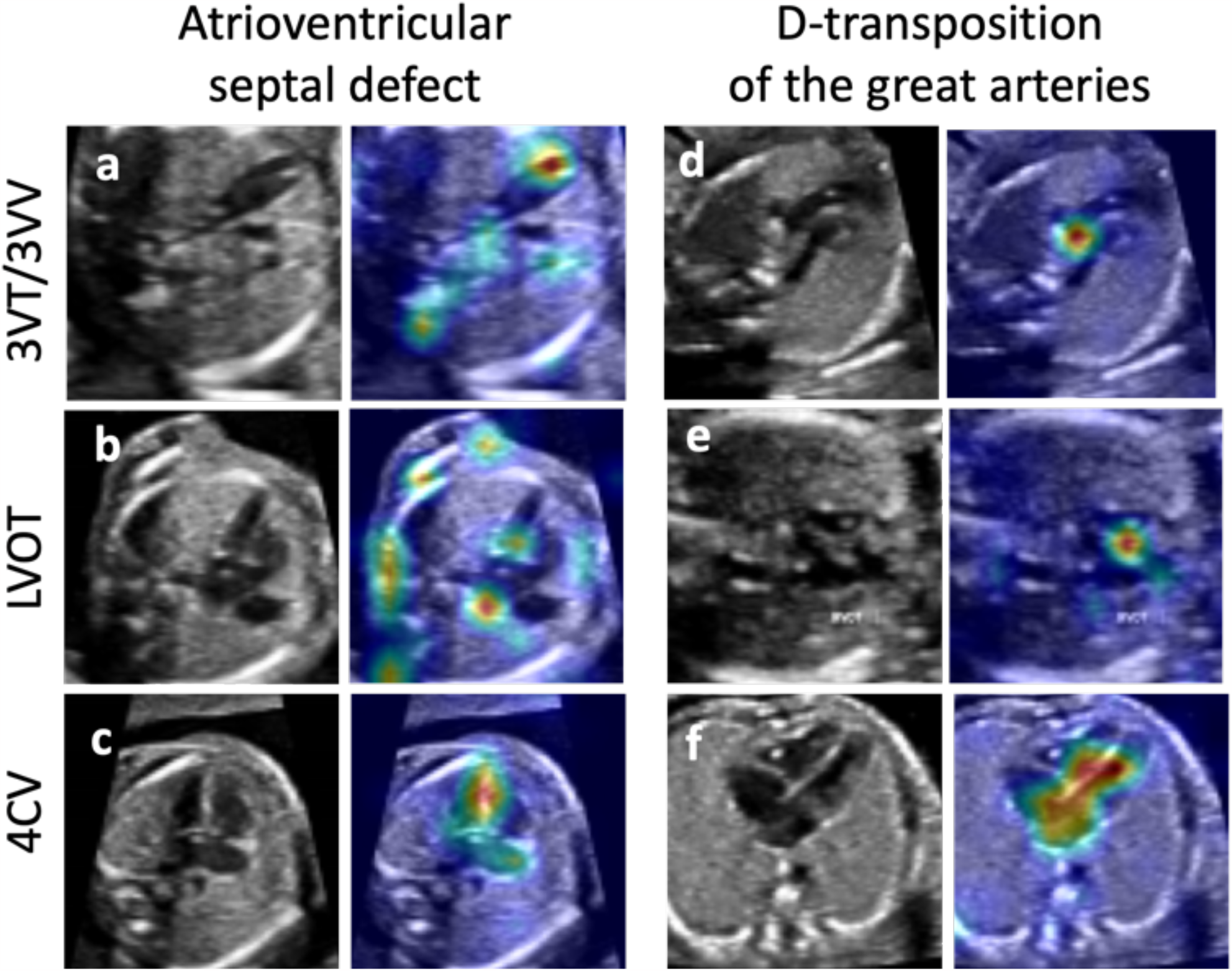
Examples of diagnostic classifier performance in studies the model got correct but blinded expert performance was variable. Model-labeled views (grayscale) with corresponding GradCAM images with corresponding GradCAM images representing a heat map showing the areas of the image most important in model decision making. Panels a-c show an example of an atrioventricular septal defect. The model was correct and all human subjects recognized the lesion. In panels d-f, an example of transposition which the model correctly identified but 2 of 3 experts incorrectly classified as normal. Abbreviations: 3VT, three-vessels-and-trachea view; 3VV, three-vessel view; 4CV, four-chamber view; LVOT, left ventricular outflow tract.

### Factors affecting ability of clinical examination, model, and retrospective blinded human expert review to correctly identify CHD

For clinical CHD detection, model, and blinded experts, we tested whether certain study features graded and described in van Nisselrooij et al were statistically different from each other based on diagnosis correctness. We report p-values for these tests in Table 3. For example, study duration in minutes was statistically different by correctness for the initial clinical CHD detection but not for the DL model nor for the blinded human experts.

**Table 3.** Difference between correct and incorrect by study feature.

| Study feature | Clinical<br>p-value<br>(MWU) | Model<br>p-value<br>(MWU) | Blinded<br>expert 1<br>p-value<br>(MWU) | Blinded<br>expert 2<br>p-value<br>(MWU) | Blinded<br>expert 3<br>p-value<br>(MWU) |
| --- | --- | --- | --- | --- | --- |
| Study duration (min) | **0.002** | 0.5 | 0.47 | 0.21 | 0.37 |
| Number of items (grayscale only, still and cine) | 0.44 | 0.44 | 0.3 | 0.36 | **0.04** |
| Number of cine items | 0.08 | 0.47 | 0.26 | 0.5 | 0.43 |
| Number of cardiac image frames (grayscale only) | 0.39 | 0.15 | 0.33 | 0.17 | **0.03** |
| Number of 3VV, LVOT, and 4CV images | 0.37 | 0.18 | 0.28 | 0.15 | **0.03** |
| Number of 4CV images | 0.13 | 0.31 | 0.29 | 0.41 | **0.02** |
| Diagnosis clear per non-blinded expert graders | **$3 \times 10^{-6}$** | 0.42 | **<0.001** | **$3 \times 10^{-6}$** | 0.25 |
| Quality score, sum of all views | **<0.001** | **0.04** | **0.01** | **<0.01** | **0.04** |
| Quality score, 3VV | 0.14 | **0.02** | 0.13 | **0.02** | **0.002** |
| Quality score, LVOT | **<0.001** | **0.01** | **0.02** | **0.02** | 0.12 |
| Quality score, 4CV | **$1.3 \times 10^{-6}$** | 0.48 | **<0.001** | 0.11 | 0.45 |
| Image quality | 0.5 | 0.31 | 0.16 | 0.17 | **<0.001** |
| CHD lesion encountered by model during training | **$1.4 \times 10^5$** | 0.07 | **0.04** | **0.03** | 0.14 |

p-values less than 0.05 are bold.

As in van Nisselrooij et al, clinical correctness also varied by quality and completeness of the cardiac screening views, namely the 4CV and LVOT view quality scores. Overall, quality and completeness of views mattered to the model and to blinded human experts as well; however, for the model and for Expert 3, quality of the 4CV view alone was not significant.

Number of frames, and presence or absence of stored cine imaging, mattered less for both the model and for the blinded human experts. Only one human improved with the presence of stored cine and greater total number of images.

Whether or not non-blinded expert grading considered the diagnosis clearly evident from stored imaging was statistically significant for clinical detection and for the blinded human experts, but not for the model.

### Cine

Though not part of the screening anatomy scan recommendations at the time of the clinical examinations, 10 of the CHD patient studies had cine captures archived. Nine of those fetuses were initially recognized to be abnormal clinically, and patients with cine were statistically more likely to be diagnosed prenatally (9/10 versus 27/57, Fisher’s exact p=0.016). The model was correct in 9/10 (90%) when cine was available, similar to its overall detection rate. On those same 10, human expert reviewers averaged only a 56% pickup rate, suggesting that cine clips may be only a surrogate for adequate information and do not necessarily contain the information themselves.

### Model detection of axial screening cardiac views

While for clinical detection and blinded human experts, view detection is implicit, for the DL model view detection is an explicit step. We compared model view detection to that of a non-blinded expert grader as a ground truth. Overall (normal and CHD hearts, all grayscale images) F-score comparing model view classification to ground truth was 0.86, representing good agreement. On normal hearts only, F-score was even higher at 0.96. (F-score for CHD hearts only was 0.85). Examples of views detected from both normal and abnormal hearts, along with their corresponding GradCAMs is shown in Figure 4.

**Figure 4.**
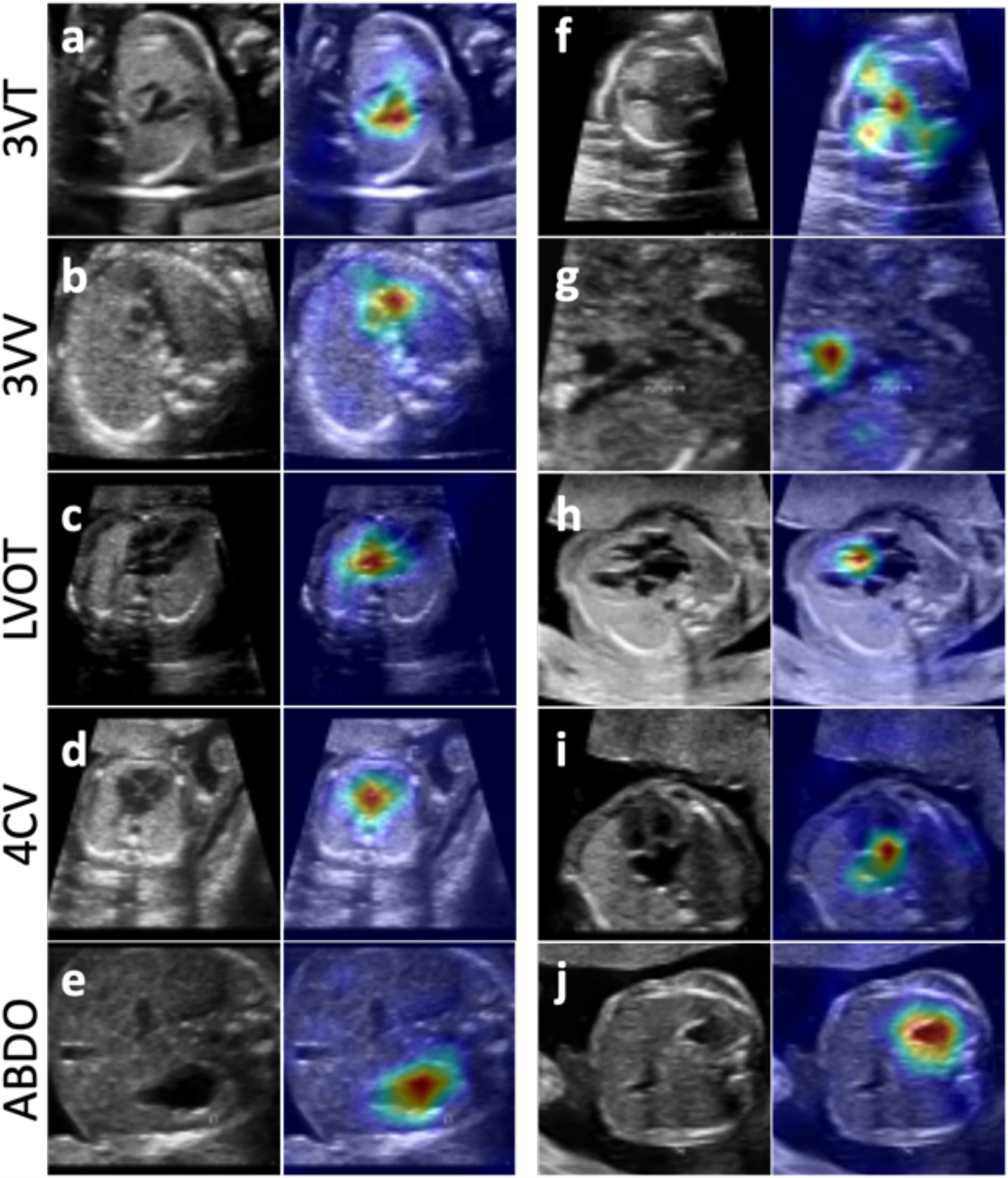
Model view finder is working and clinical features are used in model decisions. Example images with corresponding GradCAM images from view-finding step in normal (a-e) and CHD (f, d-TGA; g, Aortic stenosis; h, d-TGA, I, L-TGA; j, right atrial isomerism) cases. GradCAM images represent a heat map showing the areas of the image most important in model decision making. Abbreviations: 3VT, three-vessels-and-trachea view; 3VV, three-vessel view; 4CV, four-chamber view; ABDO, abdomen; LVOT, left ventricular outflow tract; RVOT, right ventricular outflow tract, TGA, transposition of the great arteries.

The model is compared against ground truth by the number of subjects containing a given view, as well as the average number of frames per view, in Table 4. Consistent with the F-scores described, agreement between model and ground truth is good. In addition to there being more images stored for CHD hearts than for normal hearts as mentioned above, there is a wide variability as to the number of images per view stored especially for the CHD hearts, reflecting the presence of cine in some studies as mentioned above.

**Table 4.**
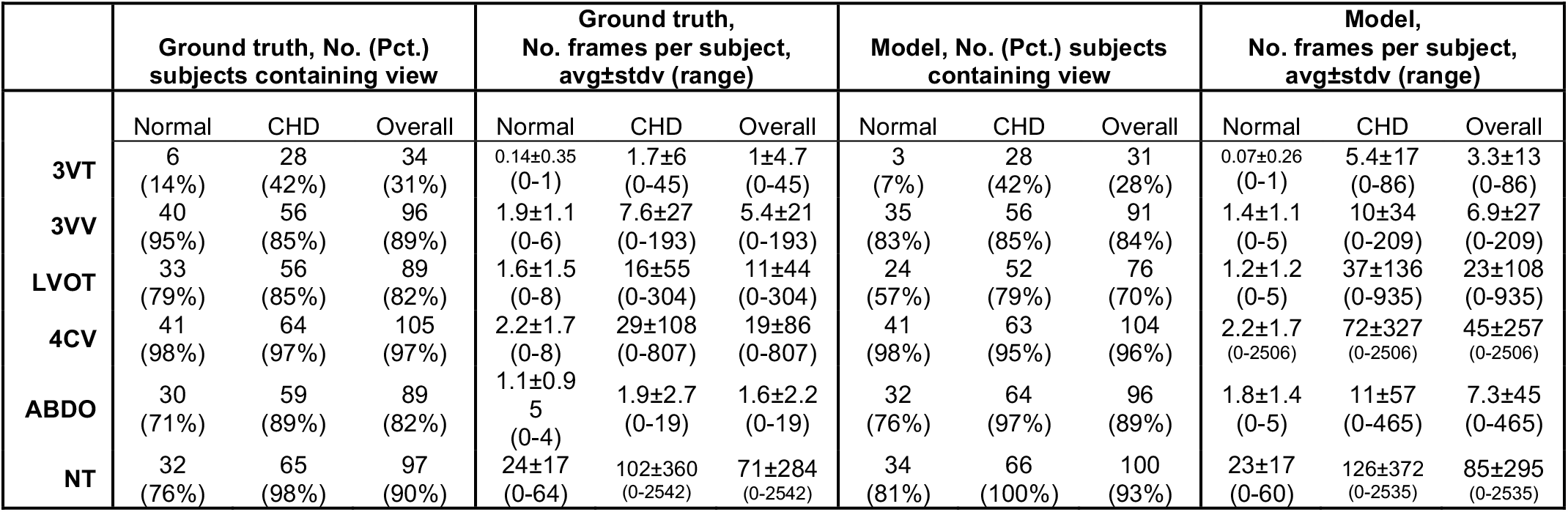
Model view detection compared to ground truth.

Finally, although the 3VT view was not part of the imaging protocol for this cohort, 3VT (ground truth) images were present in 14% of the normal studies and 42% of the CHD studies, and these were detected by the model as well (Table 4). Thus, the model can find views even when not explicitly provided by the protocol. That said, in this cohort 3VT was sufficiently rare— present in only 34 of 108 studies, and in 21 of those 34 only one 3VT image per study—that we ultimately excluded it from consideration in model diagnosis (Methods).

## Discussion

Previously, we showed that a deep learning model could be used to identify normal hearts vs. CHD from images from tertiary medical centers(10). Model design parallels clinician tasks, first finding guidelines-recommended views and then classifying those views as normal or not. In the current study, we expanded model testing using standard anatomy scans from the community— a critical step in ensuring that DL solutions are inclusive of all healthcare settings(13)—from a group of patients with known outcomes and image-by-image, view-by-view, expert-graded study quality.

Using this well-characterized community cohort, we compared model performance to both clinical detection at the point of care, as well as to the performance of human experts blinded to the study cohort’s composition and outcomes and with access to only the stored images. The model had higher sensitivity than clinical detection in this cohort, as the majority of clinically missed CHD cases were flagged by the algorithm. Importantly, expert grading found that the most substantial diagnostic errors arose from either sonographer failure to capture adequate (quality and number) images or clinician failure to recognize the abnormality from captured images. Our model presents a potential improvement less vulnerable to these obstacles.

While for simplicity the model can be said to detect normal vs. CHD, the model was not in fact trained to detect specific CHD lesions (a task already performed quite well by fetal cardiologists)(21,22). Rather, the model is designed as a screening tool to distinguish normal screening ultrasound studies from those that are *either* abnormal *or* require further review (e.g. due to incomplete or poor-quality imaging). As such, the model’s false-positive rate is high (10/42), and it cannot currently replace a trained clinician in deciding to refer patients for fetal echocardiogram. However, it may be a useful clinician aid, decreasing the number of obviously normal studies they must review, and flagging studies that are either abnormal or that could benefit from more image acquisition at the point of care.

This study has several strengths. First, the diversity of the imaging cohort(23) represents a robust test for the DL model. This cohort is external to the dataset the model was trained on, differs with respect to image formats and scanning protocol, and represents imaging from several clinics and sonographers. Number of images and cines per study are both different from the model’s training dataset and highly variable. Finally, this cohort includes a range of CHD lesions including several the model did not encounter during training. Despite this diversity, model performance is robust, compatible with the model’s suspected function as an anomaly detector, which is appropriate for screening.

A second strength is the selection of cases from a regional registry which captures missed CHD cases as well as those detected by clinicians. A third strength was including blinded humans experts to evaluate stored imaging. While clinical detection was of course an important comparator, sonographers at the point of care had access to more imaging than what was stored. Therefore, the evaluation of blinded clinical experts on stored images alone was closer to the task that the model performed.

Finally, the ability to compare clinical, model, and blinded human performance to a community imaging dataset that had been graded for quality and completeness was a strength. We found that model correctness, like that of the blinded human experts, correlated with quality measures, suggesting that the model performance is largely based on clinically relevant features. The fact that the model performed well on cases of missed CHD that were felt to be inevitable based on stored imaging is interesting: either the model detected features present in the stored imaging that are not evident to humans, or, the model was again acting as an anomaly detector.

Despite its strengths, there are weaknesses of both the current DL model and the dataset evaluated. Selection of patients for this cohort used was limited according to those who consented. One important limitation of the current model is that it can only input grayscale imaging. In the future, the model may be redesigned to accommodate color imaging.

Additional model training and algorithmic improvements may decrease its false positive rate mentioned above. However, it is worthwhile to note that the number of model false positives in this study may be at least partly attributable to a limited number of stored images. Supporting this theory is that the F-score on detected images is high. Also in support is the comparable specificity of the blinded human experts when, like the model, they lack additional imaging they would have access to in a clinical setting and are stripped of cognitive bias about the prevalence of CHD in the community.

Another model limitation is suggested by the more variable GradCAM results for the model diagnosis step as compared to the view detection step. While GradCAM is a useful way to visualize areas of the image that were important to the model in making its decision, and GradCAM heatmaps that focus on anatomical structures of clinical interest is encouraging, a ‘poor GradCAM,’ i.e. a heatmap that does not clearly focus on clinically relevant features does not necessarily mean that model performance is poor. How best to analyze GradCAMs to understand model function is still an active area of research outside the scope of this study (24– 26). Nevertheless, one imagines that an even more robustly trained model in the future may yield even better diagnostic performance.

Indeed through evaluating stored imaging in this study, we demonstrate how variable stored imaging is in number and views covered; the model didn’t have enough stored imaging to analyze two studies. While recent recommendations to store cine from screening ultrasound are helpful(27), further standardization of stored imaging through a combination of guidelines and point-of-care integration will likely improve clinical evaluation as well as computational screening.

In the future, a larger study evaluating a consecutive population of normal and CHD studies, making use of more standardized image storage and/or integration of a deep learning model into the point of care, perhaps with an updated DL model, would help move prenatal screening forward.

## Data Availability

Data sharing is not applicable to this article as no new data were created or analyzed in this study.

## Acknowledgments

CA, SR, AMG, and RA were supported by the National Institutes of Health and the Department of Defense (both to RA). AMG and RA supported by a generous grant from Georges Harik and Christine Hahn. AvN was supported by a grant from Stichting Hartekind. We thank the clinical experts who served as blinded human research subjects including Christine Springston, RDCS, and others, who wished to remain anonymous.

## Supplement

**Table S1.**
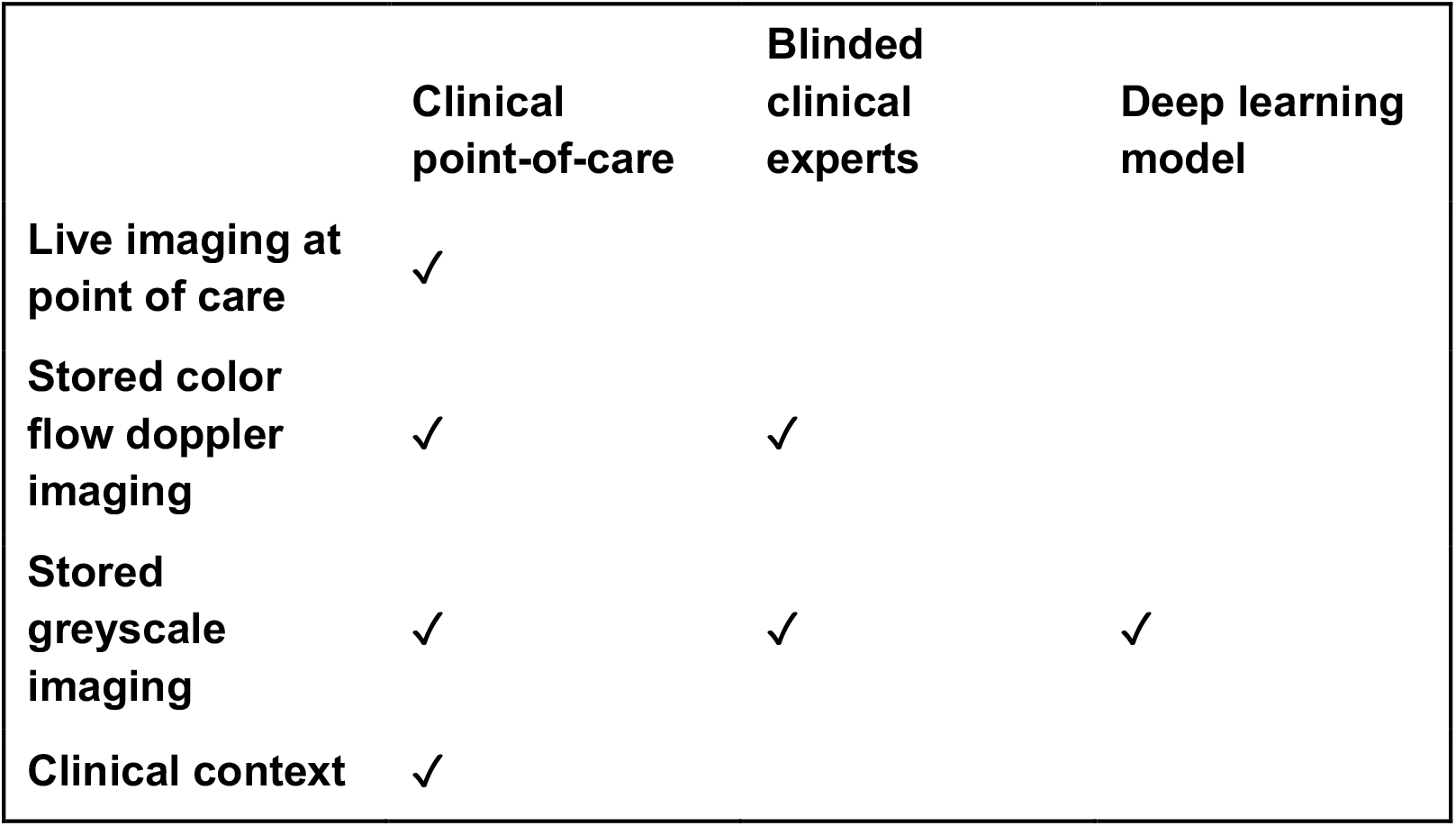
Imaging data available for clinical decision making, for blinded human experts, and for deep learning model. “Clinical context” refers to the expected prevalence of CHD in the studies being evaluated.

**Figure S1.**
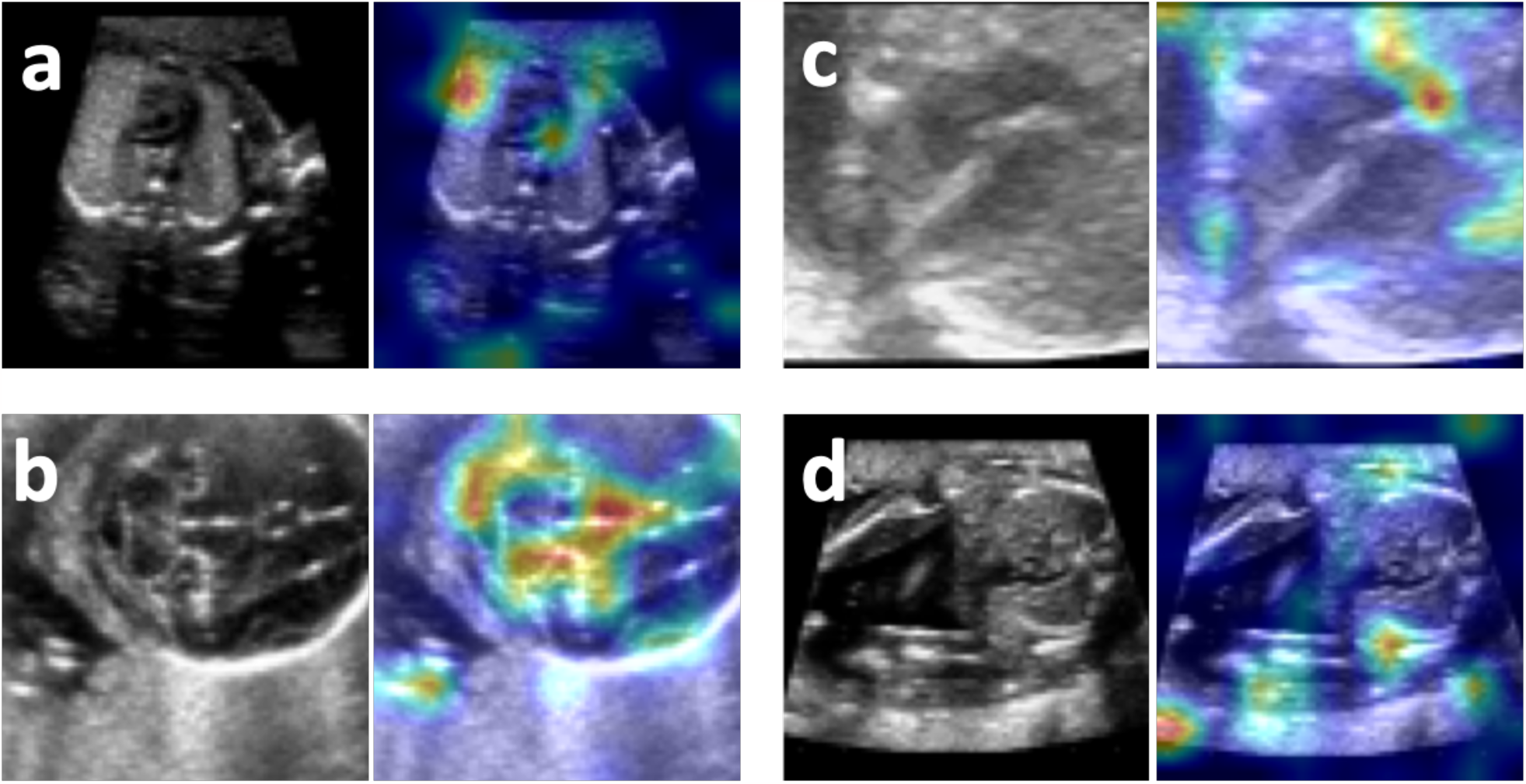
Poor model performance–errors made by model in view finding and in classification. (a) Correct view (3VV) incorrectly classified as abnormal; (b) incorrect view (LVOT) appropriately called abnormal; (c) correct view (4CV) incorrectly classified as abnormal (possibly due to poor image quality); (d) correct view (3VT) incorrectly called abnormal (possibly due to low magnification). Abbreviations: 3VT, three-vessels-and-trachea view; 3VV, three-vessel view; 4CV, four-chamber view; LVOT, left ventricular outflow tract.

